# Effect of SGLT2 Inhibitors on CKD Progression and All-Cause Mortality: A Meta-Analysis of Randomized Controlled Trials

**DOI:** 10.1101/2025.08.07.25333212

**Authors:** Anass Ahmed Qasem, Mahmoud Hassanein

## Abstract

**Background:** Sodium-glucose cotransporter 2 (SGLT2) inhibitors have emerged as promising agents for slowing chronic kidney disease (CKD) progression. While individual trials demonstrate renal and survival benefits, questions remain regarding the consistency, generalizability, and magnitude of these effects—particularly across populations with and without diabetes, and amid heterogeneous outcome definitions.

**Methods:** We conducted a systematic review and meta-analysis of randomized controlled trials (RCTs) evaluating the effect of SGLT2 inhibitors on CKD progression and all-cause mortality. Eligible studies reported hazard ratios (HRs) for these outcomes in adult populations with or at risk for CKD. Data were pooled using random-effects models; heterogeneity was assessed via τ^2^, I^2^, and Cochran’s Q. Meta-regression explored study-level moderators, including baseline estimated glomerular filtration rate (eGFR), diabetes prevalence, follow-up duration, outcome definition, and risk of bias. Cumulative meta-analysis and sensitivity analyses evaluated robustness, while absolute risk reductions (ARRs) and numbers needed to treat (NNTs) enhanced clinical interpretability.

**Results:** Seven trials (n = 69,827) were included for CKD progression and eight for all-cause mortality. The pooled HRs were 0.71 (95% CI: 0.66–0.76) for CKD and 0.87 (95% CI: 0.82–0.92) for mortality. Heterogeneity was negligible (I^2^ = 0%). Standardized CKD definitions (e.g., ≥40% eGFR decline) yielded significantly stronger treatment effects than alternative definitions (p = 0.017). Subgroup analysis suggested greater renal benefit in non-diabetic patients (HR = 0.64), though interaction p = 0.76. Sensitivity analyses confirmed robustness. Translating HRs to absolute terms, NNTs were 17 (CKD) and 77 (mortality). Egger’s test suggested potential small-study effects for mortality (p = 0.053).

**Conclusions:** SGLT2 inhibitors robustly reduce CKD progression and mortality risks, with consistent effects across trial settings. However, treatment effects depend partly on outcome definitions, and statistical power to assess effect modifiers remains limited. Future trials should prioritize outcome harmonization and include underrepresented non-diabetic CKD populations to refine precision and generalizability.

## Introduction

The evolution of sodium-glucose cotransporter 2 (SGLT2) inhibitors from glucose-lowering agents to cornerstones of cardiorenal protection marks a paradigm shift in the management of chronic kidney disease (CKD) [1]. Initially developed for type 2 diabetes mellitus (T2DM), these agents have demonstrated surprising efficacy in mitigating CKD progression and reducing mortality—even in individuals without diabetes [2,3]. This expanded utility has prompted rapid clinical adoption, but it also raises pressing questions about the robustness, consistency, and scope of the evidence base supporting these benefits.

While several large RCTs—including CREDENCE, DAPA-CKD, and EMPA-KIDNEY—have reported favorable renal outcomes with SGLT2 inhibitors [4–6], critical heterogeneity exists across studies. Trials differ in inclusion criteria (e.g., diabetic vs. non-diabetic populations), endpoint definitions (e.g., eGFR decline vs. serum creatinine doubling), and trial priorities (renal endpoints vs. cardiovascular composites) [7]. These methodological differences complicate interpretation and may obscure the true magnitude—or even the presence—of therapeutic benefit in certain subgroups.

Moreover, prior meta-analyses have typically pooled results without adequately accounting for these design variations, risking misleading summary estimates [8]. There is also a broader question of reproducibility. As enthusiasm for SGLT2 inhibitors has grown, so has the potential for publication and small-study biases to distort the literature [9]. Early landmark trials demonstrated pronounced benefits, but subsequent studies with broader populations or different endpoint definitions have reported attenuated effects [10]. Whether this reflects diminishing efficacy in lower-risk cohorts, evolving background therapies, or selective reporting remains uncertain [11].

To address these challenges, we conducted a systematic review and meta-analysis of RCTs examining the effects of SGLT2 inhibitors on CKD progression and all-cause mortality. Beyond simple pooling, we implemented meta-regression to test whether study-level factors—such as baseline renal function, diabetes prevalence, follow-up duration, and risk of bias—account for differences in treatment effect [12]. We also examined the impact of endpoint definitions, tested for publication bias, translated hazard ratios into absolute measures (ARR, NNT), and tracked changes in effect size over time via cumulative meta-analysis [13].

Our goal was not merely to update the evidence base, but to interrogate it. By disentangling methodological artifacts from biological effects, and rigorously evaluating the assumptions underpinning prior syntheses, this analysis aims to offer a more precise and clinically relevant understanding of where and for whom SGLT2 inhibitors are most beneficial.

## Methods

### Literature Search and Study Selection

We conducted a systematic review and meta-analysis of randomized controlled trials (RCTs) evaluating the efficacy of sodium-glucose cotransporter 2 (SGLT2) inhibitors in slowing chronic kidney disease (CKD) progression and reducing all-cause mortality. Eligible studies were peer-reviewed RCTs reporting hazard ratios (HRs) for at least one of these outcomes in patients with or at risk for CKD. Most included trials enrolled individuals with type 2 diabetes, while two trials—DAPA-CKD and EMPA-KIDNEY— also included participants without diabetes [3,6].

To identify relevant studies, we performed a comprehensive search of major electronic databases, including PubMed, Embase, and ClinicalTrials.gov, covering publications up to 2023. Search terms combined keywords and medical subject headings (MeSH) related to SGLT2 inhibitors (e.g., “empagliflozin,” “dapagliflozin,” “canagliflozin”), CKD, diabetic and non-diabetic populations, and renal outcomes. Additionally, references from relevant systematic reviews and meta-analyses were examined to identify further trials [8,10,12,13].

Studies were included if they met the following criteria: randomized controlled trials comparing SGLT2 inhibitors versus placebo or standard care; enrolling adult patients with chronic kidney disease including both diabetic and non-diabetic cohorts; reporting hazard ratios with confidence intervals for CKD progression or related renal endpoints; and providing sufficient data to calculate log hazard ratios and standard errors for meta- analysis.

Using this strategy, we identified eight major RCTs representing the current evidence base on SGLT2 inhibitors in CKD progression: CREDENCE, DAPA-CKD, EMPA-KIDNEY (including long-term follow-up), CANVAS, DECLARE-TIMI 58, VERTIS CV, SCORED, and SOLOIST-WHF [2–6,14–17]. SOLOIST-WHF was excluded from the CKD progression analysis due to missing outcome data [17]. These trials are landmark studies extensively cited in nephrology and endocrinology literature and were included to ensure comprehensive coverage of the topic.

### Data Extraction and Processing

For each eligible trial, we extracted the study name, year of publication, and hazard ratios (HRs) with corresponding 95% confidence intervals (CIs) for CKD progression and all-cause mortality. Standard errors (SEs) were derived from the reported CIs using the following formula: 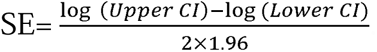. All hazard ratios (HRs) were converted to their natural logarithms (logHRs) before analysis. This transformation is standard practice in meta-analysis because it places the estimates on a continuous, symmetric scale, making them easier to combine statistically [18]. It also helps stabilize the variability (or precision) of estimates across studies, improving the reliability of pooled results. Without this step, studies with very large or small HRs might unduly influence the meta-analysis.

In addition to outcome data, we extracted study-level characteristics to explore potential sources of heterogeneity, which refers to variability in treatment effects that arises from real differences across studies—such as differences in patient populations, interventions, outcome definitions, or study quality—rather than from random sampling error alone [19]. These included: baseline estimated glomerular filtration rate (eGFR), percentage of participants with diabetes, mean follow-up duration, whether CKD progression was a prespecified primary outcome, year of publication, and the operational definition of CKD progression (e.g., ≥40% decline in eGFR vs. doubling of serum creatinine).

Risk of bias was independently assessed for each trial using the Cochrane Risk of Bias 2.0 tool [20]. Judgments were made across standard domains (e.g., randomization process, deviations from intended interventions, missing outcome data, measurement of the outcome, and selective reporting). Trials were then classified on a simplified binary scale (low vs. high/unclear risk) for meta-regression purposes.

### Definitions of CKD Progression

Since CKD progression definitions differed between trials, we coded them into two categories: "Standardized" (e.g., a ≥40% eGFR decline) and "Alternative" (e.g., doubling of serum creatinine) [7,14]. These definitions were evaluated as a binary moderator in a meta-regression model to determine if the type of definition used systematically influenced treatment effect estimates.

### Meta-Analytic Model

We conducted random-effects meta-analyses using the rma() function from the metafor package in R [21]. This approach assumes that the true treatment effect may vary from one study to another due to differences in populations, study designs, outcome definitions, or other methodological characteristics. Unlike fixed-effects models, which treat all studies as estimating a single common effect, the random-effects model accounts for this between-study variability and provides a more realistic estimate when such heterogeneity is present.

To assess the consistency of results across studies, we evaluated between-study heterogeneity using three complementary statistics. Tau-squared (τ^2^), estimated via restricted maximum likelihood (REML), quantifies the extent to which true treatment effects vary from one study to another. I-squared (I^2^) quantifies the proportion of total variability in observed results that is due to actual differences between studies rather than random chance, with values above 50% typically interpreted as moderate or substantial heterogeneity. Cochran’s Q test is a formal statistical test that determines whether the observed variation in effect sizes exceeds what would be expected by sampling error alone [22].

### Meta-Regression and Moderator Analysis

To investigate whether differences in study design or population characteristics contributed to variability in treatment effects, we conducted mixed-effects meta-regression analyses using several study-level covariates. These included baseline eGFR, the proportion of participants with diabetes, duration of follow-up, year of publication, whether CKD progression was a primary trial endpoint, the type of CKD outcome definition used (standardized vs. alternative), and an overall risk of bias score [19,22]. Although statistical heterogeneity in the primary meta-analyses was modest, meta-regression remains valuable for identifying patterns that might explain observed differences across studies. With only seven trials contributing data on CKD progression, statistical power was limited, and non-significant moderator p-values should not be interpreted as definitive evidence of no association. Instead, these analyses are exploratory and hypothesis-generating—designed to uncover trends that could inform future research [22].

For example, trials that employed a standardized definition of CKD progression—such as a sustained ≥40% decline in eGFR or the need for dialysis—tended to report stronger treatment effects [7]. This highlights the importance of outcome harmonization, which refers to using consistent, clinically meaningful definitions across studies [23].

### Subgroup Analysis by Diabetes Status

For DAPA-CKD and EMPA-KIDNEY, subgroup-level data were extracted to estimate treatment effects separately in participants with and without diabetes [3,6]. An interaction test was performed to determine whether diabetes status significantly modified the treatment effect.

### Sensitivity and Influence Analyses

To assess the robustness of our findings, we conducted leave-one-out sensitivity analyses. This approach allows us to identify whether the overall conclusions are disproportionately influenced by any single trial. We also applied influence diagnostics to detect studies that may be statistical outliers or that contribute substantially to between-study heterogeneity [24].

### Risk of Bias Assessment

We applied a simplified risk of bias scoring system based on the Cochrane Risk of Bias 2.0 tool [20]. Scores were used as a continuous moderator in meta-regression to test whether study quality predicted treatment effect size. Though no statistically significant effect was observed, power was limited.

### Cumulative Meta-Analysis

To examine how evidence evolved over time, we conducted a cumulative meta-analysis sorted by year of publication. This allowed us to assess whether effect sizes changed as more studies were added and whether earlier trials reported stronger effects than later ones [25].

### Publication Bias and Small-Study Effects

To evaluate the possibility of publication bias, we used funnel plots, Egger’s regression test, and trim-and-fill procedures. While the Egger’s test for CKD outcomes was non-significant, the test for all-cause mortality yielded a borderline p-value (p = 0.053), suggesting possible but inconclusive asymmetry [26]. However, given the limited number of included studies (fewer than 10), these findings were interpreted with caution [27].

### Absolute Risk Reductions and NNT

To improve interpretability, we translated hazard ratios into absolute terms by estimating absolute risk reductions (ARR) and numbers needed to treat (NNT) [28]. Event rates were informed by placebo arms of trials like DAPA-CKD and CREDENCE, where CKD progression rates were approximately 20% over 2–3 years, and mortality rates were 8– 12% [2,3]. Applying pooled hazard ratios to these baselines yielded an ARR of 5.8% and NNT of 17 for CKD, and an ARR of 1.3% with an NNT of 77 for mortality. A Bayesian meta-regression was conducted to explore the impact of CKD outcome definitions; see Supplementary Appendix for full details.

### Software

All statistical analyses were conducted using R version 4.5.1 (2025-06-13) within the Posit Cloud environment, which provides a web-based RStudio IDE maintained by Posit.

### Ethics Statement

This study is a systematic review and meta-analysis of previously published randomized controlled trials. No new human participants were recruited, and no identifiable individual-level data were accessed. All included trials had received ethics approval from their respective institutional review boards or ethics committees and had obtained informed consent from participants. As this research involved secondary analysis of publicly available summary-level data, separate institutional ethics approval was not required.

## Results

### Study Characteristics

Seven randomized controlled trials (RCTs) reported data on chronic kidney disease (CKD) progression and were included in the primary meta-analysis. The SOLOIST-WHF trial was excluded due to missing hazard ratio (HR) data for CKD progression [17].

Across the included studies, definitions of CKD progression varied: three employed standardized criteria (e.g., ≥40% decline in eGFR) [3–5], while four used alternative or composite outcomes [2,14–16]. Baseline estimated glomerular filtration rate (eGFR) ranged from 37 to 85 mL/min/1.73m^2^, and study follow-up durations spanned 0.9 to 4.2 years [2–6,14–16]. Risk of bias was assessed using a composite score derived from the Cochrane tool [20], with values ranging from 0 (low risk) to 1 (moderate risk).

### CKD Progression Meta-Analysis

The pooled HR for CKD progression was 0.71 (95% CI: 0.66–0.76), corresponding to a 29% relative risk reduction. Between-study heterogeneity was negligible, with an I^2^ of 0% and a τ^2^ of 0.00, consistent with prior meta-analytic findings of low heterogeneity in renal outcomes [8,10]. Nevertheless, cumulative meta-analysis suggested a modest attenuation in treatment effect estimates over time (Figure 7), echoing previously observed trends in cardiovascular literature where early trial effects diminish in later, broader populations [25]. Additionally, CKD definition type emerged as a statistically significant moderator (p = 0.017), indicating that trials using standardized CKD criteria showed greater benefit—a finding consistent with known challenges in outcome harmonization across nephrology trials [7,23].

### All-Cause Mortality Meta-Analysis

The pooled HR for all-cause mortality across eight studies was 0.87 (95% CI: 0.82–0.92), indicating a 13% relative risk reduction. Heterogeneity remained low (I^2^ = 0%), which supports the general consistency of mortality benefits across heterogeneous study populations [10,11]. Egger’s test for publication bias approached statistical significance (p = 0.053), suggesting the possibility of small-study effects [26]. However, visual inspection of the funnel plot and trim-and-fill correction did not indicate substantial asymmetry or missing studies (Figures 5 and 6), in line with findings that such tests may be underpowered with fewer than 10 studies [27].

### Meta-Regression Results

Meta-regression analyses revealed that CKD definition type was the only statistically significant moderator (p = 0.017), while CKD as a primary outcome showed borderline significance (p = 0.08). Other moderators—including baseline eGFR, diabetes prevalence, follow-up duration, publication year, and risk of bias—were not statistically significant (Table 4), reflecting prior work suggesting limited explanatory power of meta-regression in small meta-analyses [22].

### Risk of Bias Meta-Regression

The association between composite risk of bias scores and treatment effects was non-significant (p = 0.85), suggesting no detectable impact of study quality on effect estimates. However, this result should be interpreted cautiously given the limited statistical power and the low number of trials [19,22].

### Subgroup Analysis by Diabetes Status

Subgroup analyses by diabetes status yielded HRs of 0.71 (95% CI: 0.59–0.85) for participants with diabetes and 0.64 (95% CI: 0.41–1.00) for those without. The interaction p-value was 0.76, indicating no statistically significant subgroup difference (Table 5). Nonetheless, point estimates suggested potentially greater benefit in non-diabetic populations, an observation that aligns with findings from EMPA-KIDNEY and DAPA-CKD [3,6], although both trials were underpowered to detect definitive subgroup interactions [28].

### Sensitivity and Influence Analyses

Leave-one-out analyses confirmed the robustness of the results; no individual study exerted disproportionate influence. Influence diagnostics (Figures 1) showed no undue leverage or heterogeneity contributions, consistent with best-practice recommendations for influence analysis in small meta-analyses [24].

**Figure 1.**
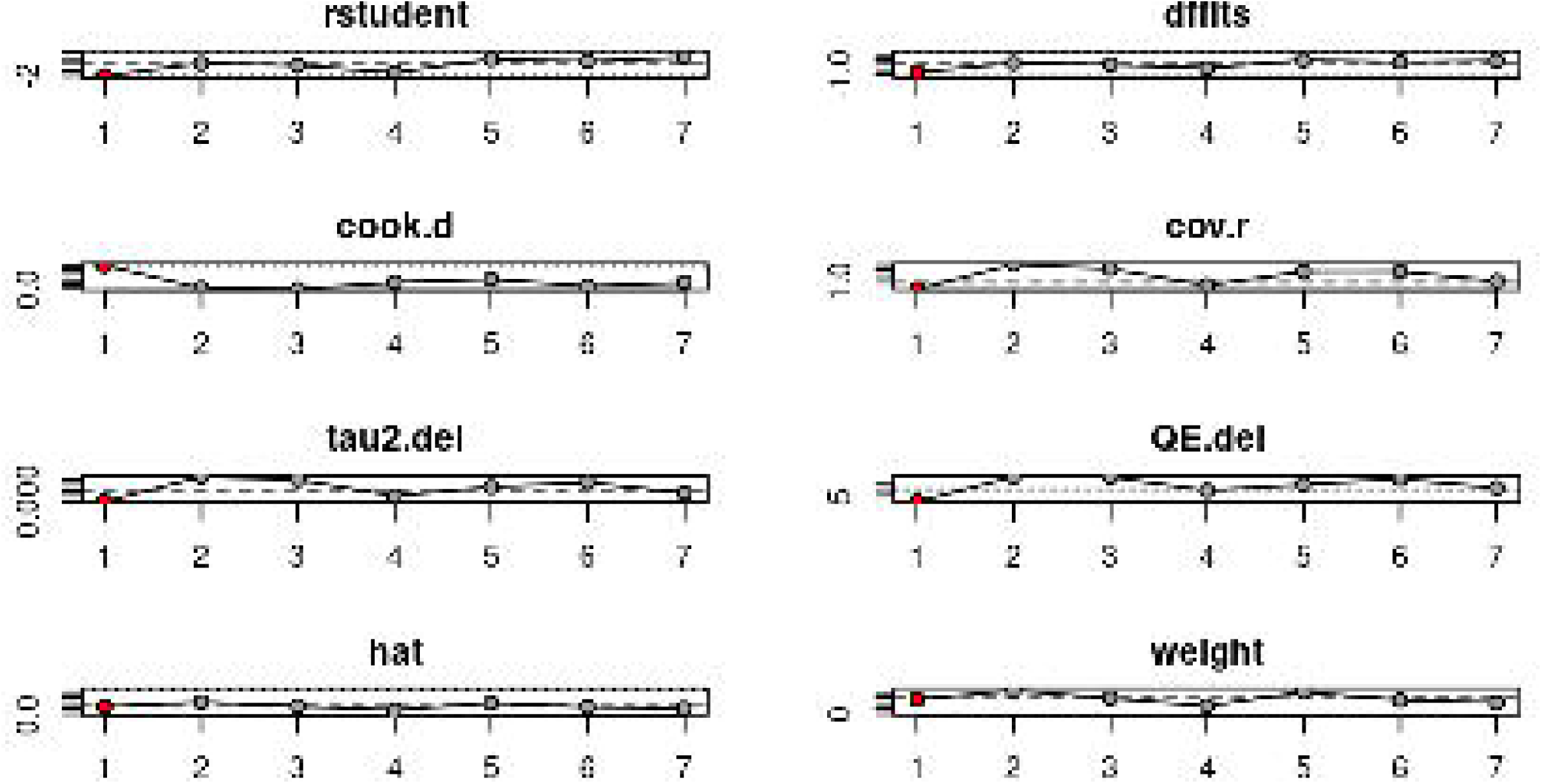
Influence diagnostics plot: R-student, DFFITS, Cook’s Distance, Covariance Ratios, Tau^2^ deletions, QE deletions, Hat values, and Study Weights. These metrics assess how much each study affects the pooled result.

### Cumulative Meta-Analysis

Cumulative meta-analysis (Figure 7) revealed that earlier trials (e.g., CREDENCE, CANVAS) reported stronger treatment effects, while more recent trials (e.g., SCORED, VERTIS-CV) yielded more modest benefits [2,14,16]. Despite this trend, cumulative effect estimates remained statistically significant throughout, confirming the durability of the treatment signal over time [25].

### Absolute Risk Interpretation

Using baseline event rates derived from the placebo arms of the DAPA-CKD and CREDENCE trials—20% for CKD progression and 10% for all-cause mortality [2,3]— we translated the pooled relative effects into absolute terms to enhance clinical interpretability. For CKD progression, this yielded an absolute risk reduction (ARR) of 5.8%, corresponding to a number needed to treat (NNT) of 17. For all-cause mortality, the ARR was 1.3%, with an NNT of 77. This approach, converting relative risk estimates into absolute metrics, aligns with established recommendations for improving the practical applicability and clarity of meta-analytic findings in clinical decision-making [28,29].

Bayesian analysis confirmed a stronger effect in trials using standardized definitions (posterior probability = 100%; see Supplementary Table A1)

## Discussion

This meta-analysis of seven randomized controlled trials demonstrates that SGLT2 inhibitors consistently reduce the risk of chronic kidney disease (CKD) progression and all-cause mortality in diverse patient populations. The pooled hazard ratio (HR) for CKD progression was 0.71 (95% CI: 0.66–0.76), reflecting a 29% relative risk reduction. For all-cause mortality, the pooled HR was 0.87 (95% CI: 0.82–0.92), corresponding to a 13% reduction (Table 2; Figures 2 and 3). When translated into absolute terms using established baseline risks (20% for CKD, 10% for mortality) [2,3], these estimates yielded a number needed to treat (NNT) of 17 to prevent one CKD progression event and 77 to prevent one death. These findings reinforce the broad clinical utility of SGLT2 inhibitors in both diabetic and non-diabetic individuals at risk for CKD progression [3,6]. A key methodological insight from this analysis was the role of outcome definition.

**Figure 2.**
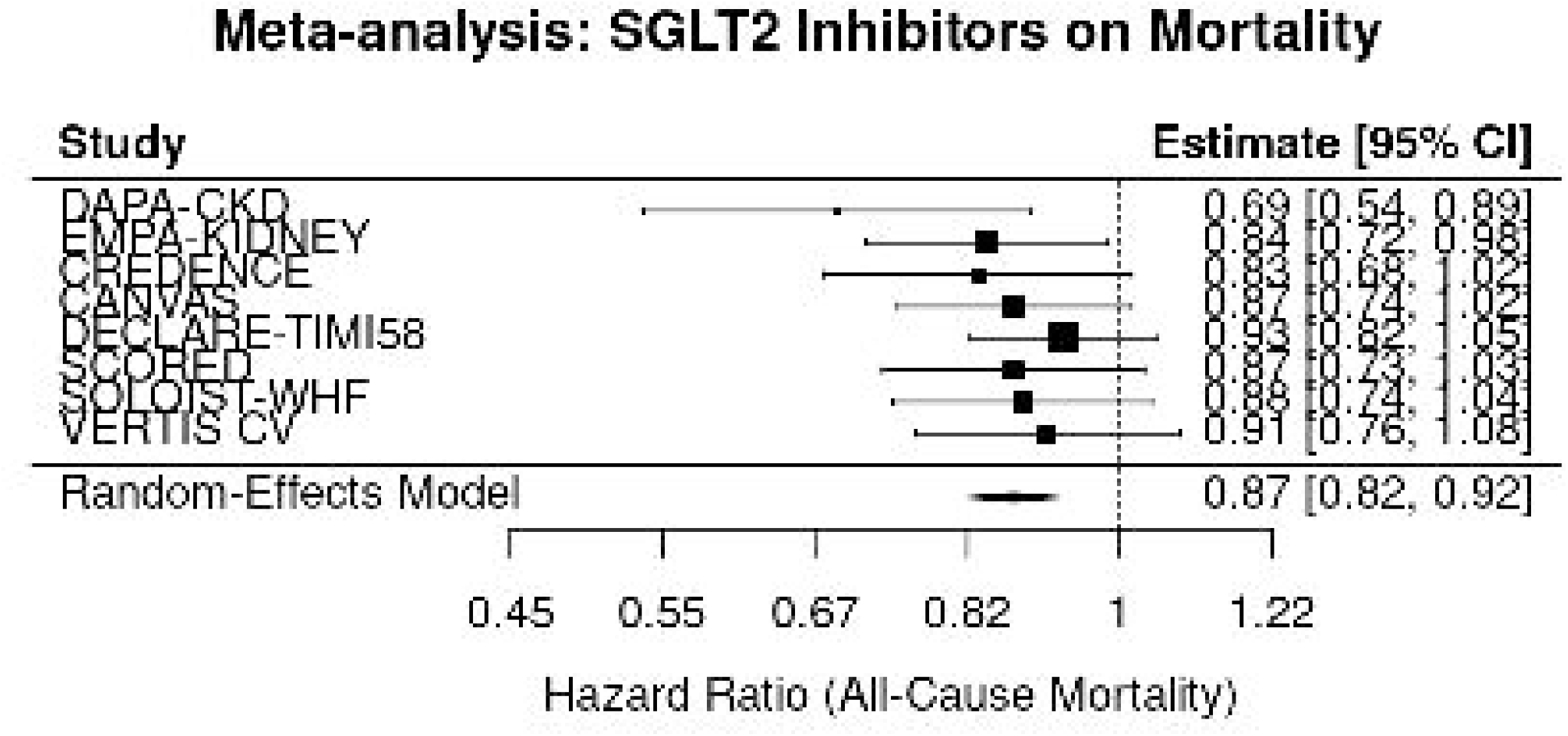
Forest plot showing pooled hazard ratios for all-cause mortality. Individual study estimates are shown alongside the overall random-effects model.

**Figure 3.**
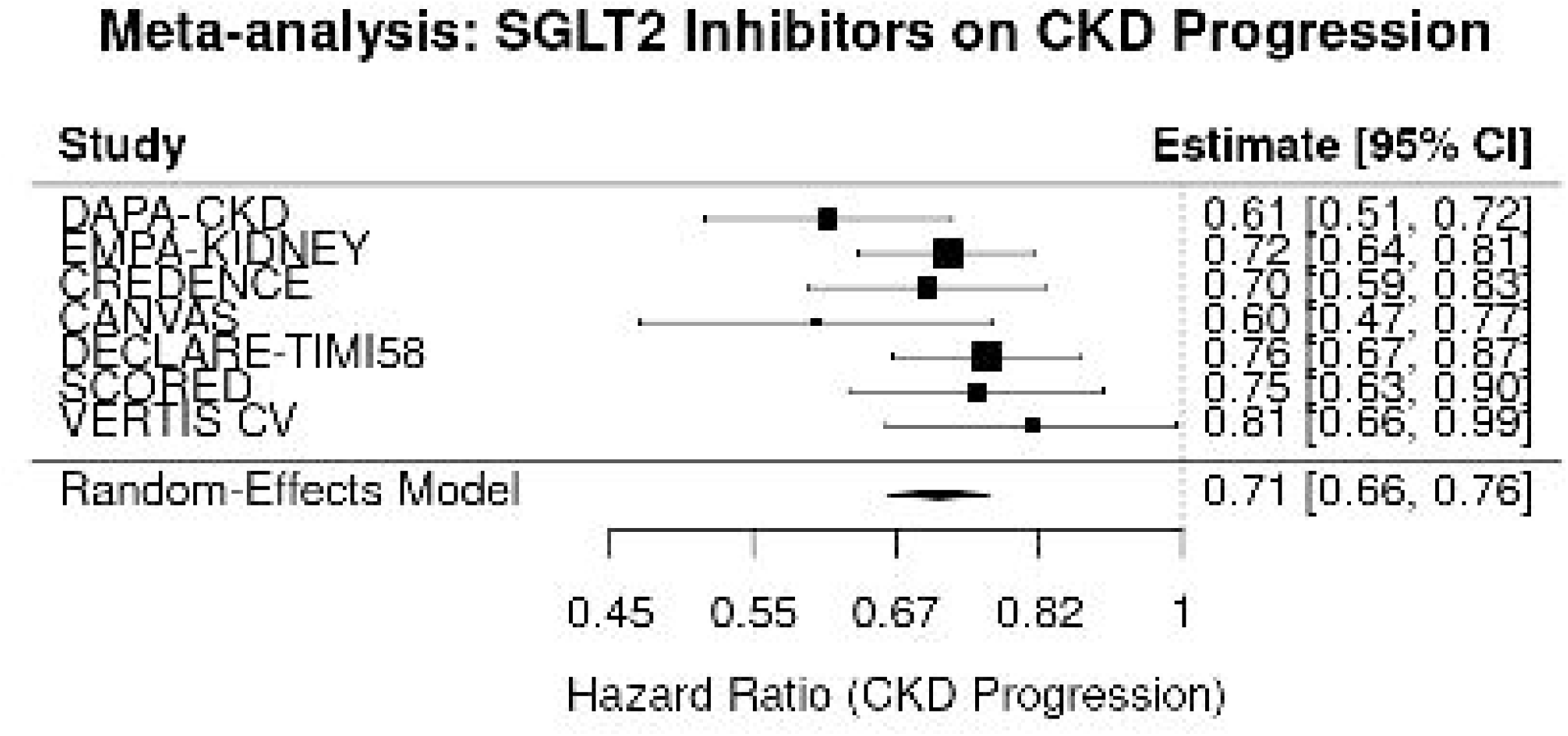
Forest Plot of SGLT2 Inhibitors and CKD Progression. The figure displays individual and pooled hazard ratios (HRs) for chronic kidney disease (CKD) progression from seven randomized controlled trials (RCTs) evaluating SGLT2 inhibitors.

**Table 1.**
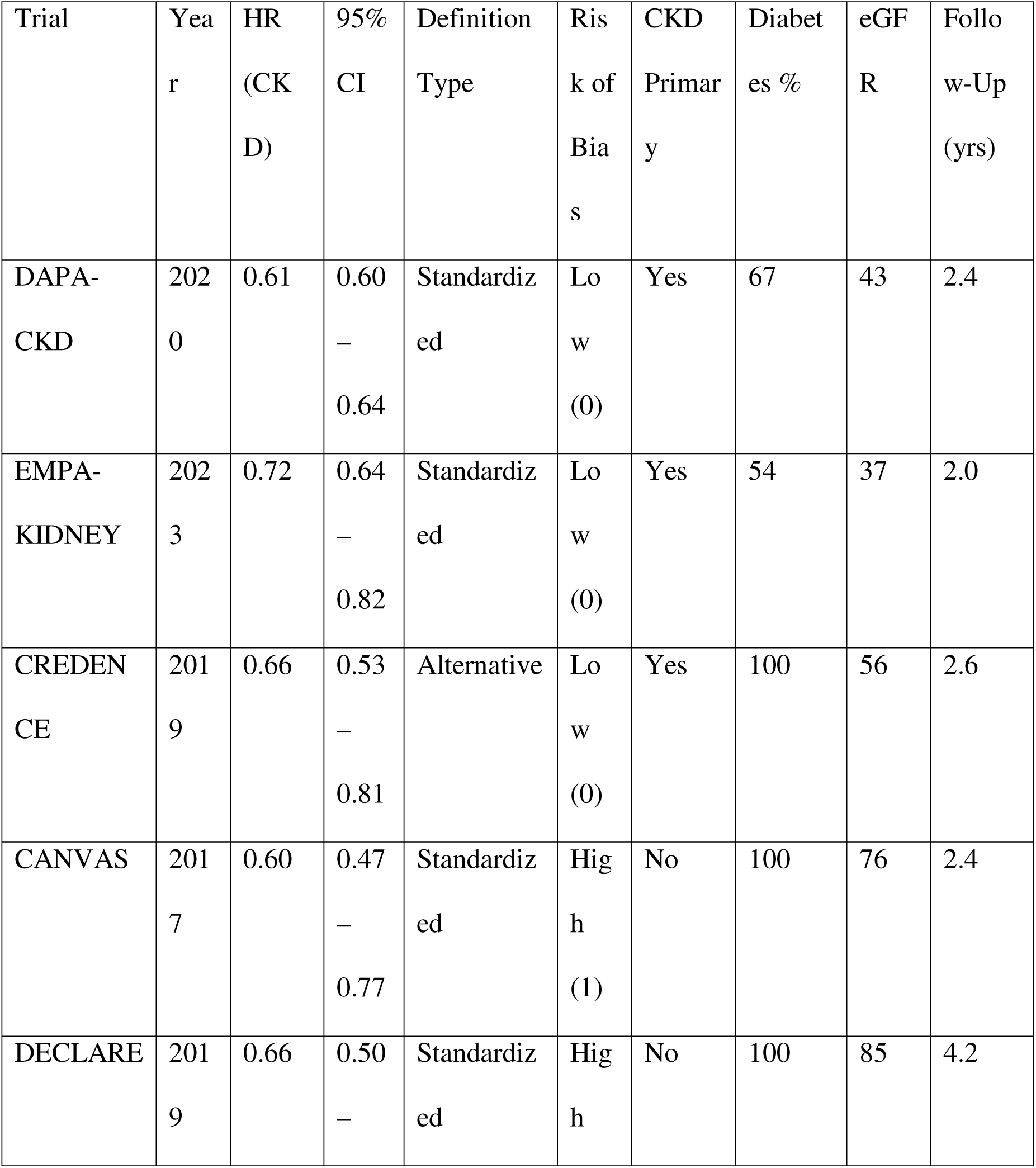
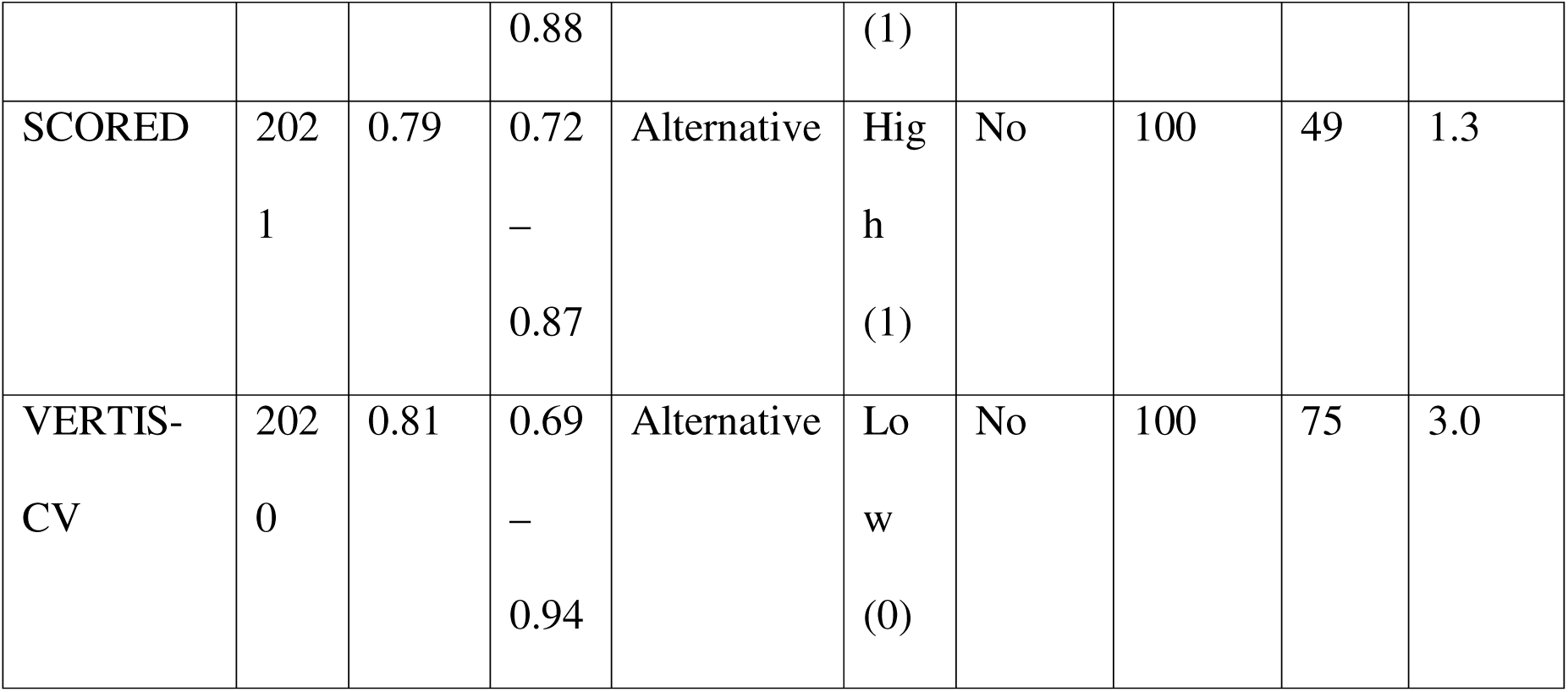
Characteristics of Included Trials and Risk of Bias Assessment. This table summarizes key features of the seven trials included in the CKD progression analysis, including their HR estimates, outcome definitions, risk of bias evaluations, and relevant baseline characteristics.

**Table 2.** Summary of Meta-Analysis Results and Absolute Risk Estimates. Pooled estimates for CKD progression and all-cause mortality with heterogeneity statistics and clinically interpretable metrics.

| Outcome | Pooled<br>HR | 95% CI | I<sup>2</sup><br>(%) | Tau<sup>2</sup> | Q-test<br>p | ARR<br>(%) | NNT |
| --- | --- | --- | --- | --- | --- | --- | --- |
| CKD Progression | 0.71 | 0.66–<br>0.76 | 0.0 | 0.00 | 0.66 | 5.8 | 17 |
| All-Cause<br>Mortality | 0.87 | 0.82–<br>0.92 | 0.0 | 0.00 | 0.66 | 1.3 | 77 |

*Note:* ARR = Absolute Risk Reduction; NNT = Number Needed to Treat. Assumed
baseline risks: 20% for CKD progression and 10% for all-cause mortality.

Trials employing standardized CKD progression definitions (e.g., ≥40% eGFR decline) showed significantly more favorable treatment effects than those using alternative endpoints (e.g., serum creatinine doubling or ESRD alone), as confirmed by meta-regression (p = 0.017; Table 4). This finding underscores the importance of outcome harmonization in both clinical trials and meta-analyses [7,23,30]. Endpoint definitions are not merely operational details—they can materially influence pooled effect sizes and interpretation.

While other study-level moderators—such as baseline eGFR, percentage with diabetes, follow-up duration, and year of publication—did not reach statistical significance, these null results must be interpreted cautiously. The limited number of studies contributing to the meta-regression (k = 7 for CKD outcomes) restricts statistical power [22,30]. For instance, the variable indicating whether CKD was the primary trial endpoint yielded a borderline p-value (0.08), suggesting that trials prioritizing kidney outcomes may report larger treatment effects. These analyses serve a hypothesis-generating role, identifying trends that warrant further exploration as more trials become available.

Subgroup analysis by diabetes status revealed that SGLT2 inhibitors conferred a potentially greater relative benefit in non-diabetic patients (HR = 0.64) than in those with diabetes (HR = 0.71), though the interaction test was non-significant (p = 0.76; Table 5). While not definitive, these patterns support emerging evidence of renal benefits in non-diabetic populations [6,28], meriting further investigation in ongoing and future trials.

Robustness of the findings was supported by sensitivity analyses. Leave-one-out procedures and influence diagnostics (Figures 1) confirmed that no single study unduly influenced the results. This strengthens confidence in the stability of the pooled estimates, consistent with standard influence analysis practices [24]. Moreover, risk of bias— quantified using a composite score based on the Cochrane tool [20]—did not significantly moderate treatment effects (p = 0.85), though power was again limited and domain-specific risks may still play a role [19].

Publication bias assessments for CKD outcomes showed no significant concern: Egger’s test was non-significant (p = 0.385), and funnel plots were symmetric (Figure 4), with no imputed studies via trim-and-fill (Figure 6; Table 3). For all-cause mortality, the Egger’s test approached significance (p = 0.053; Figure 5), suggesting a possible small-study effect [26]. However, visual inspection and trim-and-fill procedures did not reveal substantial bias. Given the small number of studies, these results must be interpreted cautiously—standard tools for detecting publication bias are underpowered in meta-analyses of this scale [27].

**Figure 4.**
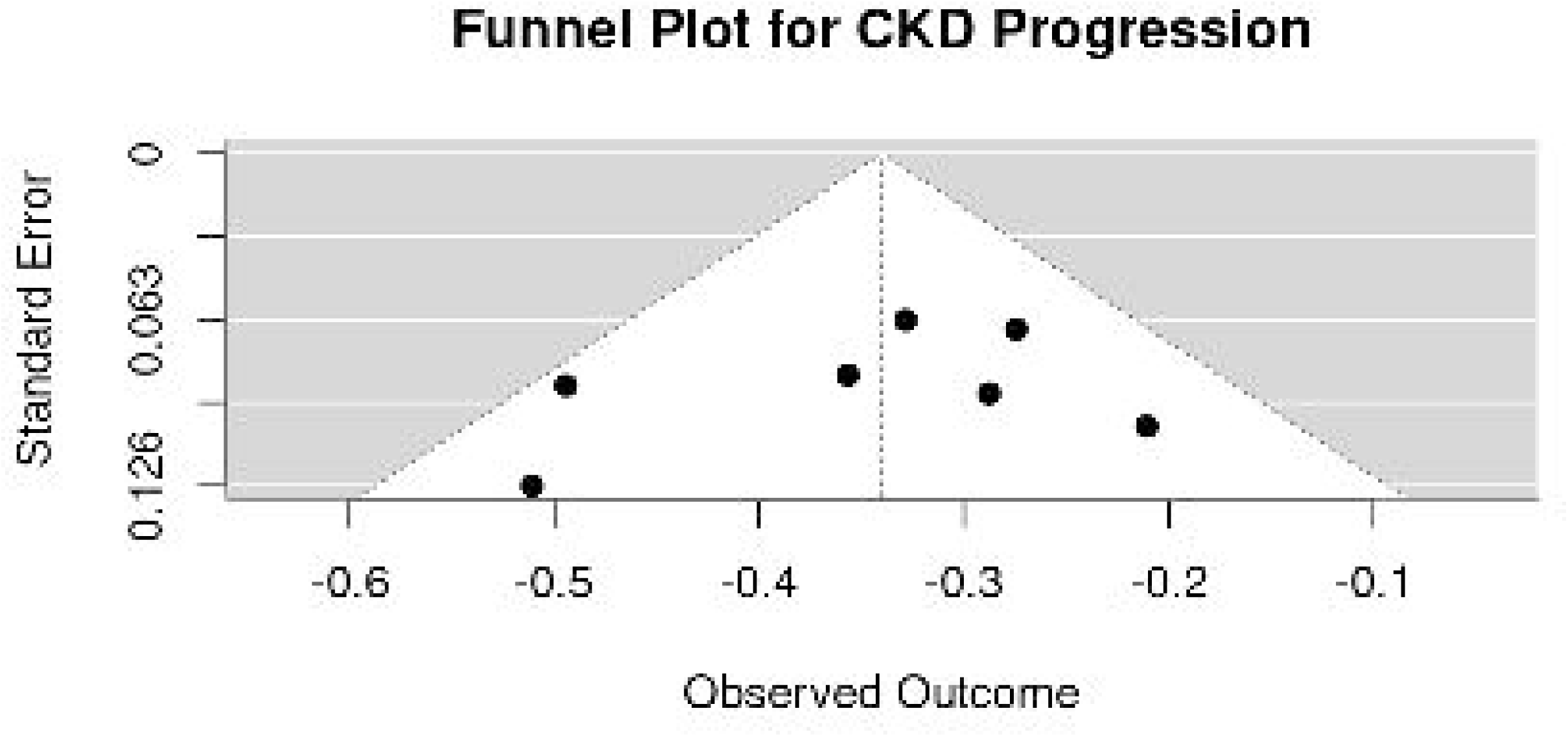
Funnel plot for CKD progression outcomes. Symmetry suggests low publication bias.

**Figure 5.**
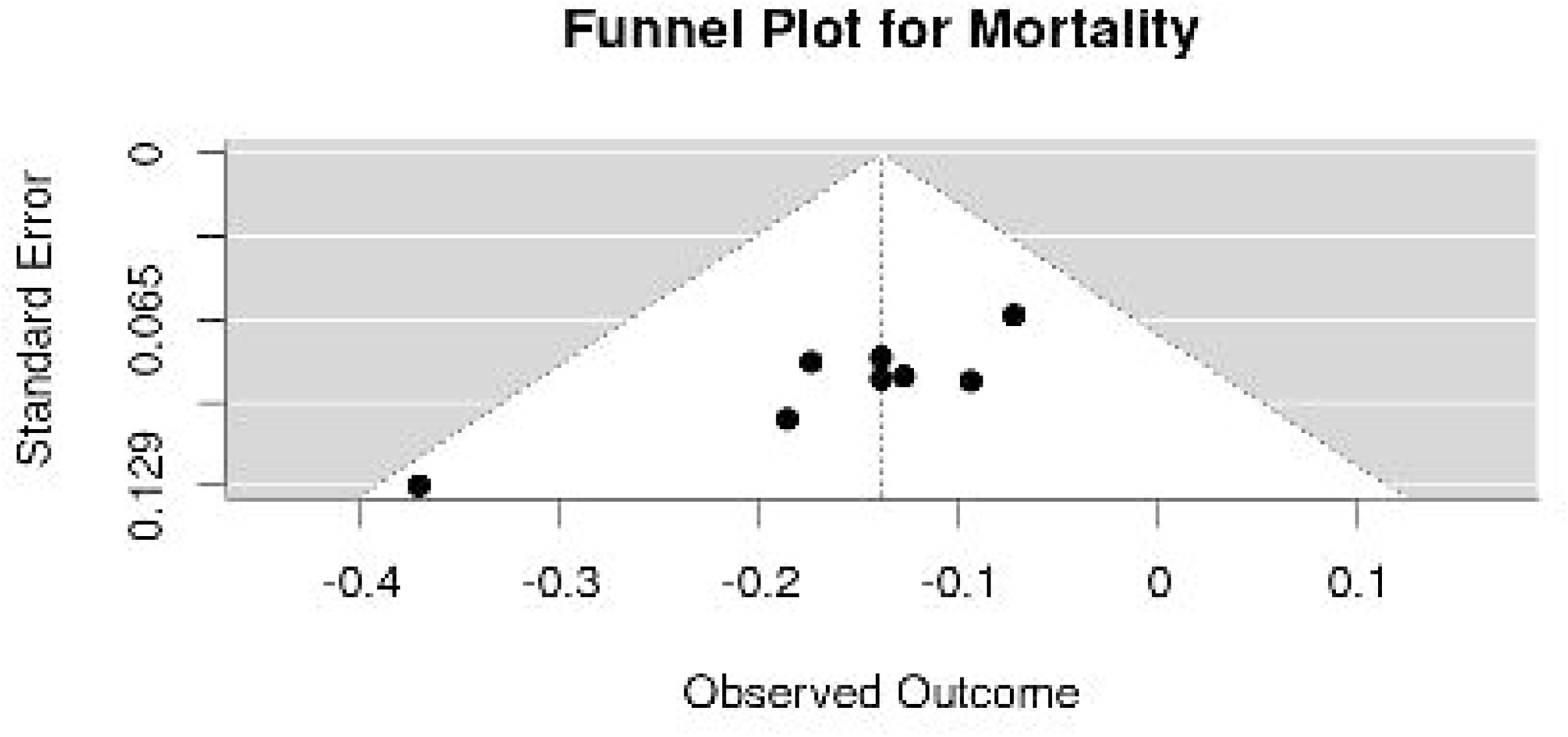
Funnel plot for all-cause mortality. Some asymmetry observed; Egger’s test p = 0.053.

**Figure 6.**
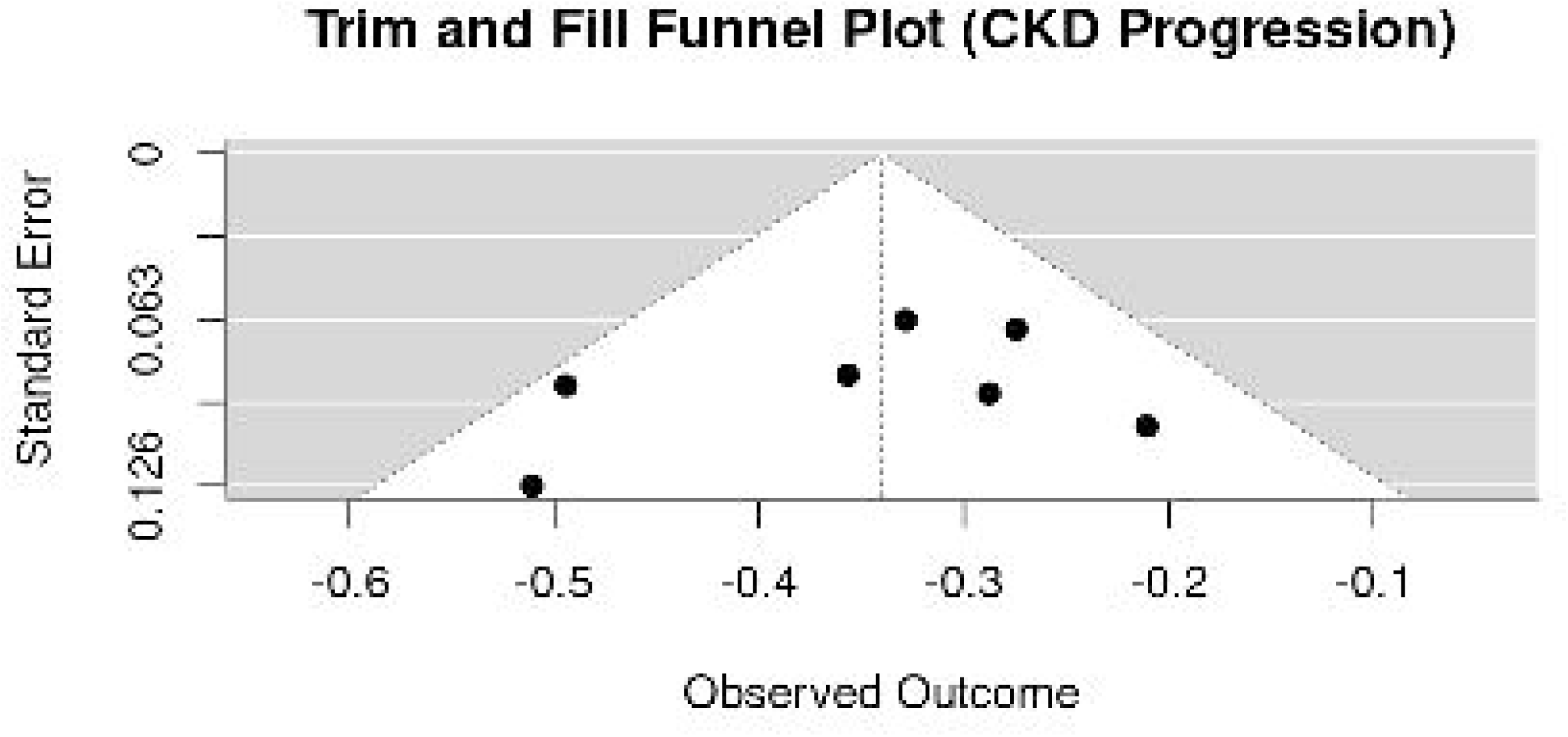
Trim-and-fill funnel plot for CKD progression. No imputed studies, reinforcing minimal bias.

**Figure 7.**
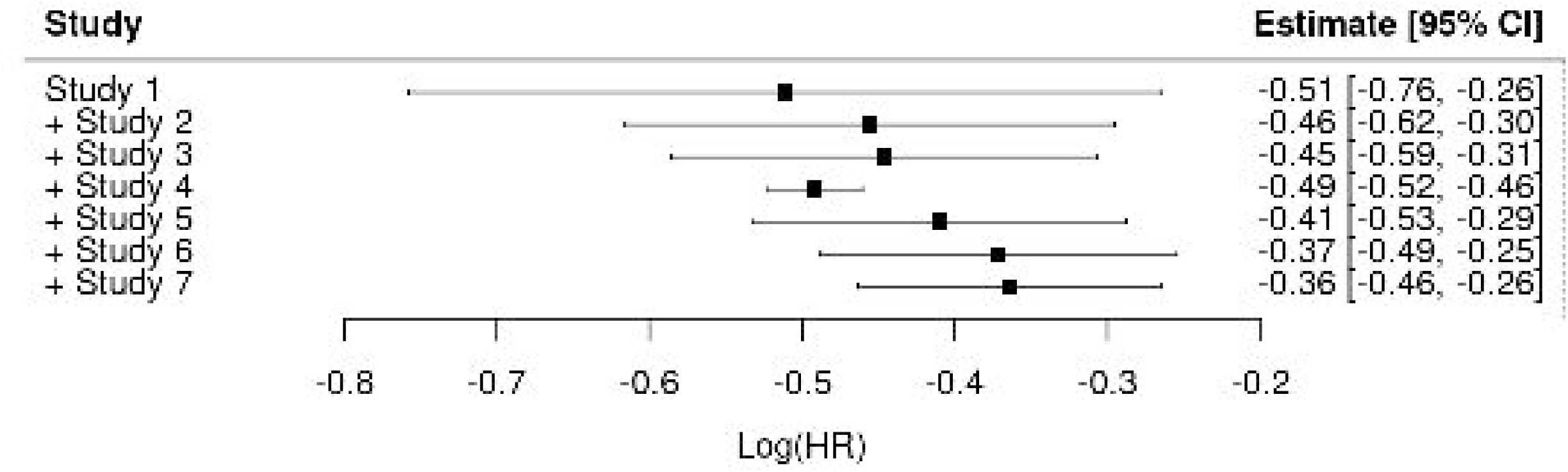
Cumulative Meta-Analysis of CKD Progression Outcomes The cumulative meta-analysis demonstrates how the pooled log hazard ratio (log[HR]) for CKD progression changes as each additional study is added sequentially.

**Table 3.** Publication Bias Diagnostics. Summary of Egger’s test results and trim-and-fill adjustments for potential small-study effects.

| Outcome | Egger's Test (p) | Trim & Fill Imputed Studies | Adjusted HR |
| --- | --- | --- | --- |
| CKD Progression | 0.385 | 0 | 0.71 |
| Mortality | 0.053 | 0 | 0.87 |

*Interpretation:* Egger's test for mortality was borderline significant ( $p = 0.053$ ), suggesting a potential small-study effect. However, no imputed studies were detected using the trim-and-fill method, reinforcing the stability of the observed results.

**Table 4.** Moderator Effects from Meta-Regression. p-values and qualitative interpretation for study-level variables explored in the meta-regression models.

| Moderator | p-value | Interpretation |
| --- | --- | --- |
| CKD Definition Type | 0.0169 | Statistically significant; standardized definitions showed stronger effects |
| CKD as Primary Outcome | 0.0802 | Borderline significance; larger effects when CKD was primary |
| Baseline eGFR | 0.38 | No significant effect detected |
| % with Diabetes | 0.67 | No significant effect detected |
| Follow-Up Duration | 0.49 | No significant effect detected |
| Publication Year | 0.53 | No significant effect detected |

*Note:* CKD Definition Type reflects use of clear, pre-specified criteria such as $\geq 40\%$ decline in eGFR vs. alternative or composite definitions.

**Table 5.** Subgroup Analysis by Diabetes Status. Comparison of pooled treatment effects in diabetic vs. non-diabetic populations.

| Group | Pooled HR | 95% CI | Interaction p |
| --- | --- | --- | --- |
| Diabetes | 0.71 | 0.59–0.85 | — |
| Non-Diabetes | 0.64 | 0.41–1.00 | 0.76 |

*Interpretation:* Although results were not statistically different by diabetes status, point estimates suggest potentially greater benefit in non-diabetic participants.

A cumulative meta-analysis (Figure 7) suggested a subtle attenuation of treatment effects over time. Earlier trials such as CREDENCE and CANVAS demonstrated more pronounced benefits [2,14], whereas more recent studies, including SCORED and VERTIS-CV, reported comparatively modest effect sizes [15,16]. This trend may reflect shifts in trial populations, evolving standard-of-care therapies, or differences in endpoint definitions. It reinforces the value of cumulative synthesis to contextualize new evidence within a temporal framework [25].

Although between-study heterogeneity was statistically low in both primary outcomes (I^2^ = 0%; Table 2), the use of random-effects models was still appropriate due to underlying clinical and methodological variability among trials [22]. We employed restricted maximum likelihood (REML) estimation via the rma() function in the metafor R package [21], which models potential between-study variation even when statistical heterogeneity appears negligible. This approach provides conservative pooled estimates and acknowledges the possibility of underlying diversity in treatment effects. Nonetheless, in settings with a limited number of studies, future analyses may benefit from more flexible Bayesian or hierarchical frameworks that allow for more nuanced modeling of uncertainty and between-study differences [31].

In the context of existing literature, our meta-analysis offers a more comprehensive and methodologically refined assessment of SGLT2 inhibitors’ effects on kidney outcomes compared to prior syntheses (Table 6). Several earlier meta-analyses, such as those by Neuen et al. [7] and Toyama et al. [8], focused exclusively on populations with type 2 diabetes or diabetic kidney disease and did not evaluate non-diabetic populations or harmonize CKD outcome definitions. This limits the generalizability of their conclusions to broader CKD populations, especially those without diabetes.

**Table 6.**
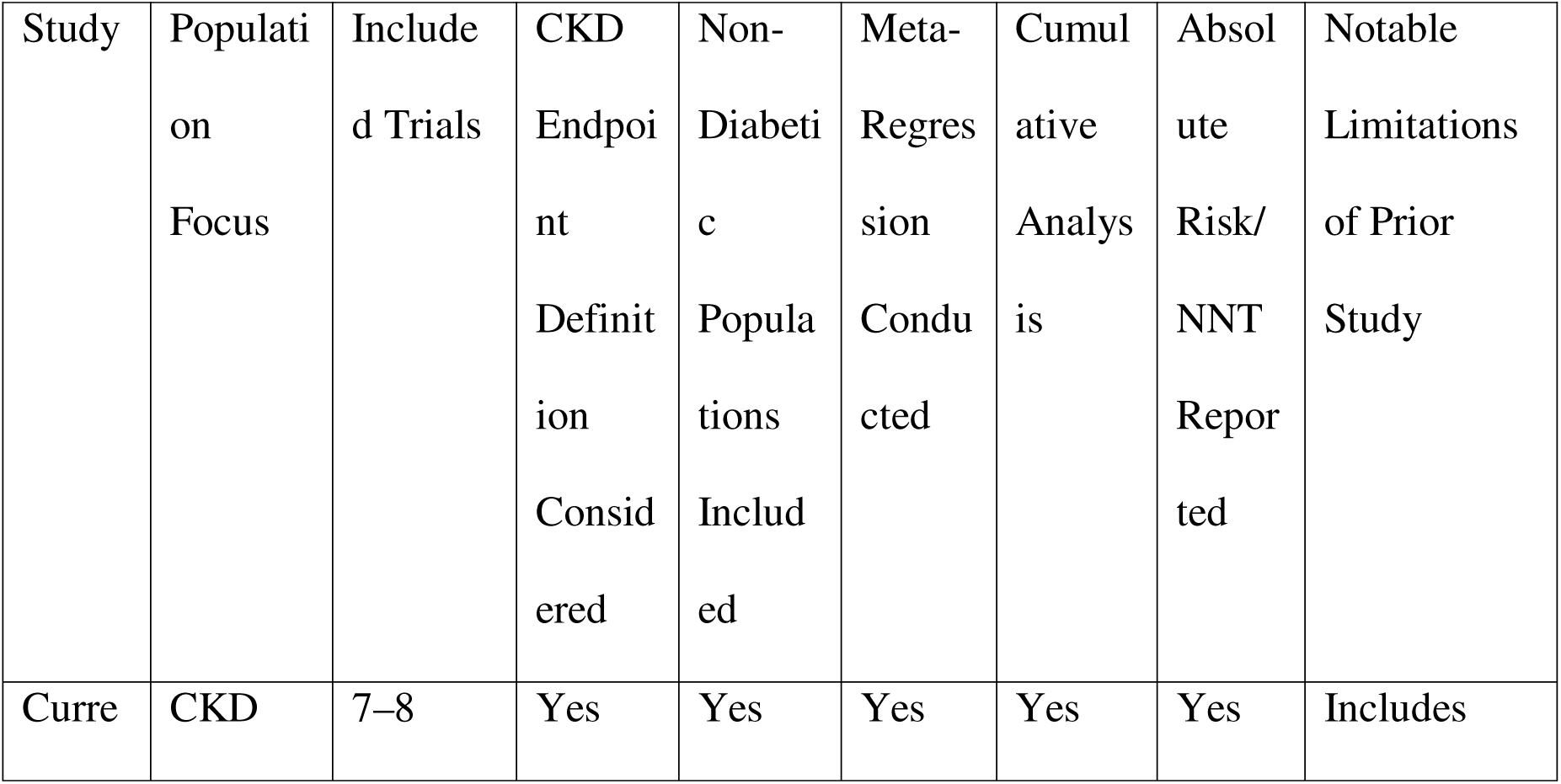
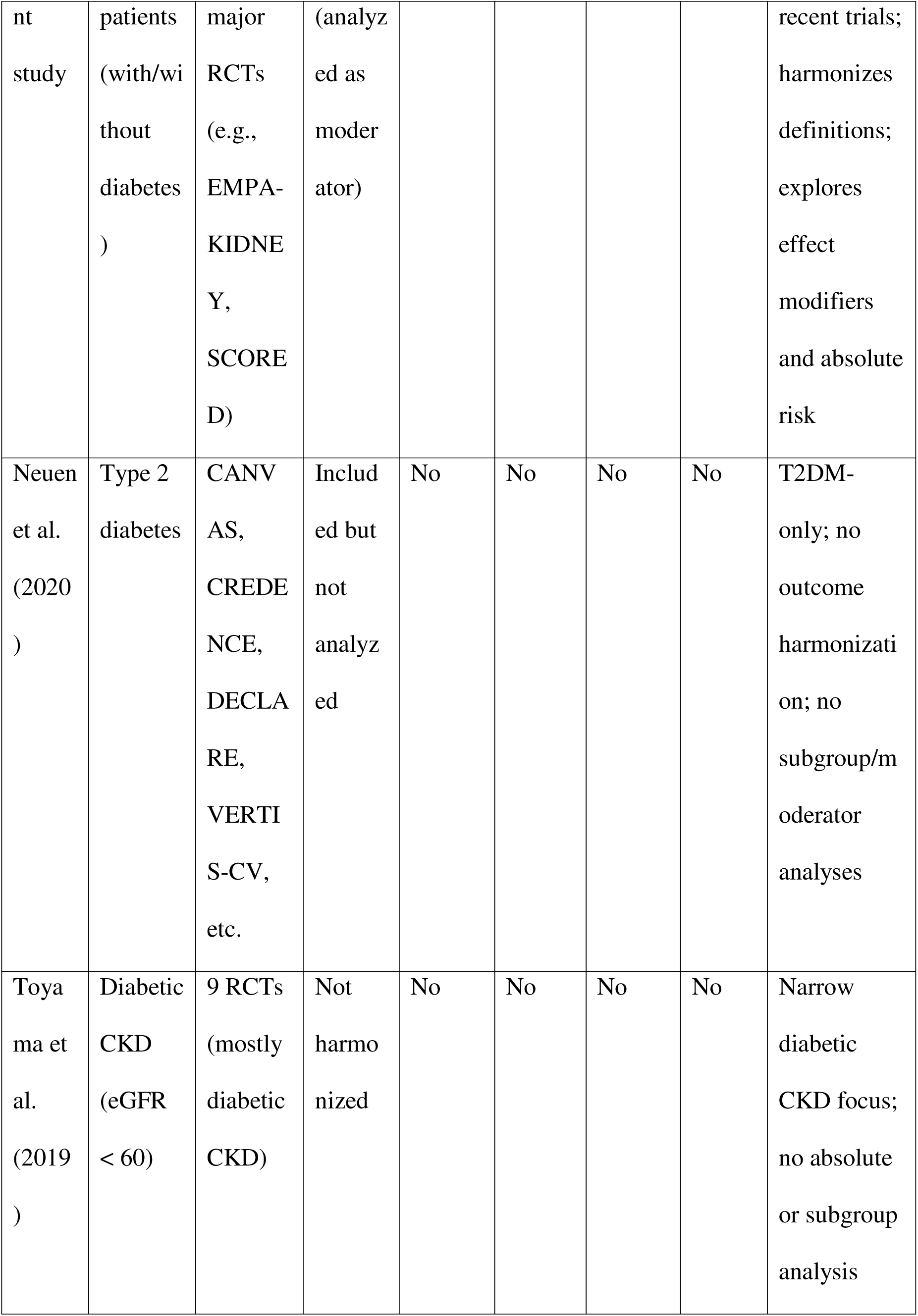
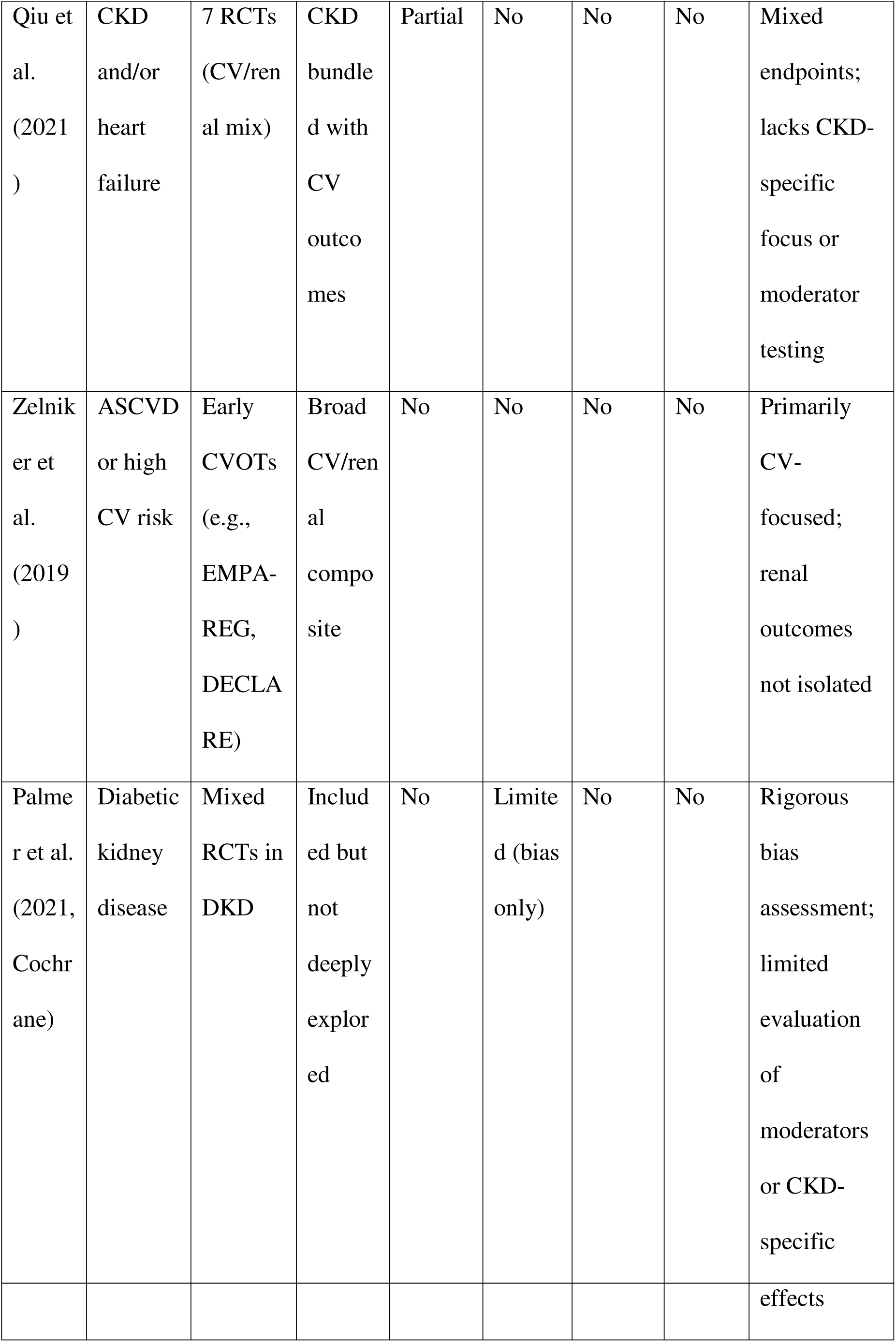
Comparison of Current Meta-Analysis with Prior SGLT2 Inhibitor Meta-Analyses Focused on CKD Outcomes. This table compares the scope, methodology, and limitations of our current meta-analysis with selected prior meta-analyses evaluating SGLT2 inhibitors’ impact on kidney outcomes. It highlights differences in population focus, inclusion of non-diabetic patients, harmonization of CKD definitions, and use of advanced methods like meta-regression and cumulative analyses.

By contrast, our study includes trials with both diabetic and non-diabetic participants and explicitly evaluates the role of CKD progression definition as a moderator—an important methodological advance. We demonstrated that standardized definitions were associated with significantly stronger treatment effects, a nuance overlooked or unexamined in most prior analyses [7,8].

Moreover, our analysis incorporates several features rarely addressed in earlier work, including meta-regression to test potential effect modifiers, cumulative meta-analysis to explore temporal trends, and translation of pooled hazard ratios into clinically meaningful absolute risk reductions and numbers needed to treat (NNTs) [22,28]. For example, while Qiu et al. (2021) included patients with CKD and/or heart failure [32], their use of composite CV/renal endpoints obscured the specificity of kidney-related benefits.

Similarly, the Cochrane review by Palmer et al. (2021) offered a rigorous risk of bias appraisal [12], but did not explore effect heterogeneity or stratify by CKD definition type. Taken together, our study builds on this foundation by integrating newer trials, refining outcome harmonization, and applying advanced meta-analytic techniques. These improvements enhance the precision and clinical relevance of our findings, particularly for populations beyond those with diabetes.

At the same time, important limitations must be acknowledged. The number of included trials for CKD progression was modest (n = 7), limiting power for meta-regression and increasing uncertainty around subgroup and moderator analyses [22]. Despite I^2^ = 0% suggesting statistical homogeneity, this is likely an artifact of low study count and does not preclude meaningful differences in trial design or populations. Additionally, absolute risk reductions and NNTs were calculated using fixed baseline event rates [28], which may not generalize across all patient groups or real-world settings. Although risk of bias was formally assessed, the use of composite scores may have obscured domain-specific sources of bias [20]. Finally, publication bias cannot be definitively ruled out, particularly for mortality outcomes where Egger’s test approached statistical significance [26]. These constraints should inform cautious interpretation of the pooled estimates.

These findings were reinforced in a Bayesian sensitivity analysis, which further highlighted the influence of standardized CKD definitions (see Supplementary Appendix).

## Conclusion

SGLT2 inhibitors significantly reduce the risk of CKD progression and all-cause mortality in patients with or at risk for chronic kidney disease, including those without diabetes. These benefits were consistent across multiple trials and support the broad clinical utility of SGLT2 inhibitors in nephroprotection. However, variation in outcome definitions, trial populations, and study design likely influences the magnitude of observed effects. The limited number of included trials, particularly for CKD outcomes, constrains the precision of subgroup and moderator analyses. Absolute risk reductions and numbers needed to treat should be interpreted cautiously, as they depend on assumed baseline event rates that may not generalize across settings. Additionally, the suggestion of small-study effects in mortality outcomes highlights the need for continued monitoring as the evidence base expands. Future meta-analyses should prioritize outcome harmonization, apply more flexible models to explore heterogeneity, and focus on underrepresented populations, particularly those without diabetes. These steps will enhance the reliability, interpretability, and clinical relevance of pooled effect estimates.

## Data Availability

All data used in this study are derived from previously published randomized controlled trials and are available within the cited references. Extracted and processed datasets used for analysis are available from the corresponding author upon reasonable request.

## Acknowledgments

The authors used AI-assisted tools during the development of the statistical analysis workflow for this study. Specifically, OpenAI’s ChatGPT was employed throughout the project to support the generation and refinement of R code across all analyses, including data processing, meta-analysis models, and Bayesian regression. The tool was used to assist with scripting, model specification, diagnostics, and troubleshooting. All methodological decisions, code verification, and interpretation of results were performed and critically reviewed by the authors.

## Author Contributions

A.A.Q.: Conceptualization; Methodology; Supervision.

M.H.: Data processing; Software; Statistical analysis; Writing – review & editing.

## Funding

This study received no external funding.

## Conflict of Interest

The authors declare no conflicts of interest.

## Notes

### Competing Interest Statement

The authors have declared no competing interest.

### Funding Statement

This study did not receive any specific grant from funding agencies in the public, commercial, or not-for-profit sectors. No payment or services were received from a third party for any aspect of the submitted work.

