## Supplementary Appendix for "Effect of SGLT2 Inhibitors on CKD Progression and All-Cause Mortality: A Meta-Analysis of Randomized Controlled Trials"

### Supplementary Appendix: Bayesian Meta-Regression of SGLT2 Inhibitor Effects by CKD Definition

This appendix provides a full account of the Bayesian meta-regression model used to assess the effect of CKD definition standardization on treatment estimates. It was conducted to complement the primary frequentist meta-analysis. The main findings of this analysis are summarized in Table A1 below.

**METHODS: Bayesian Meta-Regression**

To build on our earlier frequentist random-effects meta-analysis, we conducted a Bayesian fixed-effects meta-regression to assess whether the definition of CKD progression used in each study influenced the observed treatment effect of SGLT2 inhibitors. Bayesian methods are particularly useful in settings with limited data — in this case, seven trials — as they provide a full probability distribution for each parameter, allowing for more intuitive quantification of uncertainty.

We analyzed the treatment effect as the log hazard ratio (logHR) for CKD progression, with a binary moderator variable indicating whether a trial used a standardized CKD definition (e.g., a sustained ≥40% decline in eGFR) or an alternative one (e.g., doubling of serum creatinine or reaching ESRD). The model was fit using the brms package in R, which interfaces with the Stan probabilistic programming language to perform Bayesian estimation via Markov Chain Monte Carlo (MCMC) sampling.

We specified weakly informative priors for the model's parameters. Specifically, we used a normal distribution centered at 0 with a standard deviation of 0.3 for both the intercept and the coefficient for the CKD definition variable. This prior reflects a reasonable belief, before observing the data, that the treatment effect is likely close to null (logHR = 0, or HR = 1), while still allowing for moderate treatment effects in either direction (approximately corresponding to HRs ranging from 0.74 to 1.35). These priors help stabilize estimation by discouraging implausibly large effects without strongly influencing the results.

We calculated standard errors for each study's logHR based on their reported 95% confidence intervals. These were incorporated into the model using the se() function in brms. Because this approach accounts for the uncertainty around each study's estimate, we did not include an additional term to estimate between-study heterogeneity (τ), and residual variance (σ) was effectively set to zero.

An initial attempt to fit a random-effects Bayesian model failed due to poor convergence and divergent transitions — both indicators that the model was too complex relative to the data. In Bayesian terms, this reflects overparameterization: the model was trying to estimate more parameters than the data could reliably support. Therefore, we proceeded with a fixed-effects model, which was more stable and produced interpretable posterior estimates.

The model was run with four MCMC chains, each generating 4,000 iterations, with the first 2,000 iterations of each chain used as warm-up. The warm-up phase (also called burn-in) allows the sampler to adapt to the geometry of the posterior distribution before drawing samples for inference. This yielded a total of 8,000 post-warmup draws used for analysis. Convergence diagnostics were satisfactory, with R-hat values of 1.00 and high effective sample sizes, and no divergent transitions.

Each draw from the posterior distribution represents a plausible set of values for the model parameters — namely, the average logHR for studies using alternative CKD definitions and the additional effect seen in studies using standardized definitions. Together, these samples describe a full distribution of possible treatment effects, allowing us to estimate 95% credible intervals and posterior probabilities (e.g., the probability that the treatment effect is stronger when standardized definitions are used).

In addition to examining the posterior estimates, we formally compared models with and without the CKD definition variable using a Bayes Factor (BF). This approach quantifies how much more likely the data are under the model that includes CKD definition compared to one that does not. It provides a direct measure of evidence strength for the importance of outcome definition.

**RESULTS: Posterior Estimates and Interpretation**

In Bayesian analysis, the model combines what we believed before seeing the data (the prior) with the actual observed data (the likelihood) to produce a posterior distribution. This posterior represents our updated beliefs about the true treatment effects. Each draw from this posterior is one plausible set of values for the treatment effects, given both the data and the assumptions we started with. In this case, we are sampling from the joint posterior distribution of the average log hazard ratio for trials using alternative CKD definitions and the additional effect observed in trials using standardized definitions.

The Bayesian model estimated a log hazard ratio (logHR) of –0.25 (95% credible interval: –0.33 to –0.18) for trials using alternative CKD progression definitions, which corresponds to a hazard ratio (HR) of approximately 0.78. This indicates a 22% relative reduction in CKD progression risk with SGLT2 inhibitors under those definitions.

For trials using standardized definitions, the model estimated an additional logHR of –0.23 (95% credible interval: –0.31 to –0.15). When combined with the baseline logHR, this yields an overall logHR of –0.48 — equivalent to an HR of about 0.62, or a 38% reduction in risk.

We also calculated the posterior probability that trials using standardized definitions observed stronger treatment effects (i.e., a more negative logHR). This probability was estimated at 100%, meaning every draw from the posterior supported this conclusion. While this suggests very high confidence in the direction of the effect, we also quantified the strength of evidence using a Bayes Factor.

The Bayes Factor comparing the model with CKD definition to a model without it was approximately 1,000,000. This means the data were a million times more likely under the model that included CKD definition. By conventional interpretation standards, this constitutes extreme evidence for a moderator effect of outcome definition on treatment efficacy.

**DISCUSSION: Interpretation and Context**

This Bayesian analysis supports and extends the findings of our frequentist meta-analysis. It confirms that SGLT2 inhibitors are associated with a reduction in CKD progression and shows that the estimated treatment effect is stronger in studies that used standardized outcome definitions. The posterior probability that standardized definitions yield stronger effects was 100%, and the Bayes Factor provided formal, decisive evidence for this conclusion.

The use of a weakly informative prior helped stabilize the estimates without dominating the results, and the fixed-effects approach avoided overfitting in a small dataset. While the model does not account for residual heterogeneity, the direct inclusion of study-specific standard errors provided sufficient structure to yield interpretable results.

These findings reinforce the importance of consistency in outcome definitions across trials. The stronger observed benefit in studies using standardized definitions may reflect more reliable event capture, better alignment with regulatory endpoints, or improved clinical relevance. In contrast, variability in endpoint definitions may dilute true treatment effects and increase noise in meta-analytic comparisons.

While the Bayes Factor indicates extremely strong support for the importance of CKD definition type, the analysis remains conditional on model assumptions — particularly the fixed-effects structure and small sample size. These results should therefore be interpreted as highly suggestive, but not definitive, pending larger-scale data or hierarchical Bayesian models.

Overall, this Bayesian extension demonstrates how analytic choices — including endpoint definitions and model specification — can shape the interpretation of treatment effects. The findings add weight to the argument that harmonizing CKD progression endpoints may not only improve trial quality, but also clarify evidence syntheses and strengthen clinical guidance.

**Table A1. Bayesian Estimates of Treatment Effect by CKD Definition**

The following table summarizes the posterior estimates for treatment effects by CKD outcome definition:

| Parameter | Estimate | 95% Credible Interval | Interpretation |
|---|---|---|---|
| Intercept (Alternative) | -0.25 | [-0.33, -0.18] | LogHR for studies using alternative CKD definitions |
| CKD_defstandardized | -0.23 | [-0.31, -0.15] | Additional benefit in studies using standardized definitions |
| HR (Alternative) | 0.78 | — | 22% relative risk reduction |
| HR (Standardized) | 0.62 | — | 38% relative risk reduction |

LogHR = log hazard ratio; HR = hazard ratio. Lower values indicate stronger treatment effects. Credible intervals represent the 95% most plausible range of values for each estimate, given the data and the priors.
